# Disability in Multiple Sclerosis is Associated with Vascular Factors: An Ultrasound study

**DOI:** 10.1101/2022.09.11.22279820

**Authors:** Merlisa Kemp, Clint Johannes, Susan J van Rensburg, Martin Kidd, Ferial Isaacs, Maritha J Kotze, Penelope Engel-Hills

**Affiliations:** Medical Imaging and Therapeutic Sciences, Faculty of Health and Wellness Sciences, Cape Peninsula University of Technology, Cape Town, South Africa; Internal Medicine, Faculty of Medicine and Health Sciences, Stellenbosch University, Cape Town, South Africa; Chemical Pathology, Department of Pathology, Faculty of Medicine and Health Sciences, Stellenbosch University, Cape Town, South Africa; Data Analysis, Centre for Statistical Consultation, Department of Statistics and Actuarial Sciences, Stellenbosch University, Stellenbosch, South Africa; Chemical Pathology, Department of Pathology, Faculty of Medicine and Health Sciences, Stellenbosch University and National Health Laboratory Service, Cape Town, South Africa

**Keywords:** multiple sclerosis, disability, vascular ultrasound, lifestyle factors

## Abstract

**Background:** Although multiple sclerosis (MS) is an immune-related disorder, pharmaceutical interventions targeting the immune system do not stop or reverse disability progression; the major challenge for this condition. Studies show that disability progression in MS is associated with vascular comorbidity and brain volume loss, indicating that a multi-targeted approach is required to prevent debilitation. The aim of the present study was to examine the associations between vascular ultrasound, disability, biochemistry and lifestyle data in people with MS (pwMS).

**Methods:** Extracranial vascular ultrasound was performed on 51 pwMS and 25 age-matched controls. Sonographic interrogation determined carotid intima-media thickness (cIMT) and abnormal blood flow patterns. Disability was assessed using the Expanded Disability Status Scale (EDSS). Biochemical and lifestyle data were obtained for all participants.

**Results:** The EDSS had a highly significant positive association with the cIMT of the right (r = 0.63; p = 0.001) and left (r = 0.49; p = 0.001) common carotid arteries and negative associations with the peak systolic blood flow velocity of the right vertebral artery (r = -0.42; p = 0.01) as well as end-diastolic velocity of the left internal carotid artery (r = -0.47; p = 0.01). These associations were significantly influenced by biochemical and lifestyle factors. Both cIMT and age showed significant associations with the EDSS. When cIMT was adjusted for age in a regression analysis, the association between the EDSS and the cIMT remained significant (p < 0.01), while the age association was reduced to being significant only at 10% (p = 0.06). There was no association between the use of MS medication and the EDSS (p = 0.56).

**Conclusion:** PwMS who had increased cIMT, a surrogate marker for atherosclerosis, and reduced carotid artery blood flow velocities were at risk for greater disability over and above the effect of aging. These findings provide important information for disease management and disability prevention in pwMS. Modification of diet and lifestyle may promote the unhindered flow of essential nutritional factors into the brain in pwMS.

## Introduction

Multiple sclerosis (MS) is the most common inflammatory disorder of the central nervous system affecting young adults worldwide. MS disability is related to the death of oligodendrocytes, the cells that synthesize myelin in the central nervous system (CNS), causing dysfunction of the fatty sheaths surrounding the nerve axons with consequent disruption of signal transmission from the brain to peripheral organs and, together with neuronal death, contributes to loss of brain volume ^[1].^ Patches of dead oligodendrocytes visible as white matter lesions on magnetic resonance imaging (MRI), a hallmark of MS, occur predominantly in brain areas that are prone to oxygen deficiency, ^[2]^ suggesting that lesion formation is closely linked to oxygen delivery to the CNS by blood vessels.

Disability may be transient due to the restoration of the myelin sheaths by stem cells, the oligodendrocyte precursor cells (OPCs) residing in the CNS, emphasizing the importance of understanding what the OPCs require to fulfil this function. Managing MS disability requires more than medicines, as outlined previously by van Rensburg et al. ^[1,3].^ People with MS (pwMS) need to be empowered to challenge the present narrative that predicts inevitable progressive disability following an MS diagnosis. Many studies have emphasized the role of lifestyle factors such as diet, exercise, smoking cessation and control of hypertension to prevent disability and restore function ^[4].^ This knowledge is not always implemented into MS management.

The connection between the vascular system and MS disability has not received the necessary attention by clinicians, yet the connection is central to the management of MS and in particular the disabilities faced by pwMS. This connection is likely to be related to the fact that the survival of oligodendrocytes and myelin production by these cells are dependent on the continual delivery of oxygen and nutrients and removal of toxic waste products by the blood ^[2].^ Previous studies have investigated four main vascular abnormalities associated with MS: (i) oxygen deficiency in brain tissues (hypoxia) ^[2]^, ii) failure of the blood brain barrier (BBB) to protect the brain parenchyma against infiltration of metabolites such as fibrin(ogen) ^[5]^, (iii) decreased cerebral blood flow ^[2]^ and (iv) impaired removal of toxic waste products from the brain through the jugular veins ^[6]^. Interestingly, the impaired perfusion experienced by pwMS leads to the formation of secondary arterial vessels as a compensatory measure to enhance blood delivery to hypoxic brain tissues ^[6,7]^. Furthermore, hypertension contributes to adverse outcomes in pwMS ^[8]^. Systolic blood pressure differences of greater than 5 mm Hg between right and left arm measurements can indicate whether people are at risk for vascular abnormalities ^[9]^. Lorefice et al. ^[10]^ showed that vascular comorbidities, including hypertension, were significantly associated with brain atrophy in MS, which is the most important determinant of disability ^[1,3]^.

Ultrasound assessment of the neck vessels, including measurement of the carotid intima-media thickness (cIMT) and blood flow velocities, provides a non-invasive method to determine the extent of vascular pathology, which infers atherosclerotic factors such as the deposition of cholesterol and immune activation that impede the transfer of oxygen and nutrients further along the arteries in the brain tissue. The measurement of cIMT can therefore be employed in clinical practice as a surrogate marker for atherosclerosis ^[11]^. Since vascular health is known to be influenced by both non-modifiable risk factors such as age ^[12]^, as well as modifiable factors such as diet and lifestyle, the present study provided an opportunity for testing which of these factors had the strongest association with MS disability, as assessed by the Expanded Disability Status Scale (EDSS) ^[13]^.

The study included 51 pwMS and 25 age-matched controls without neurological disorders. Ultrasound assessment included the application of grey-scale imaging, colour, and spectral Doppler analysis to investigate the association between carotid artery disease and disability status. In order to assess whether ultrasound parameters were associated with disability, statistical comparisons were made between EDSS values, cIMT and blood flow parameters in the pwMS. cIMT is significantly related to age ^[12]^; therefore, the relative effect of age was determined in the association between the cIMT and the EDSS using a regression summary for EDSS adjusted for cIMT and age. The role of cardiovascular-related biochemical biomarkers (cholesterol, homocysteine, C-reactive protein, vitamin D, serum folate and vitamin B12) and physical activity on vascular health was investigated by determining associations with the ultrasound parameters.

## Materials & Methods

### Ethics

This investigation was a collaborative study between two universities in the Western Cape, South Africa. Ethical approval was granted by the Faculty of Health and Wellness Sciences Research Ethics Committee (FREC) (Reference no: CPUT/HW-REC 2017/H4) and the Human Research Ethics Committee (HREC) of the University of Stellenbosch (Reference no: N07/09/203). The study was conducted according to the code of ethics of the World Medical Association ^[14]^ and study participants gave signed informed consent for their data to be used. Ultrasound examinations of these participants were performed, by permission, at a private radiology practice.

### Study participants

A total of 48 females and three males with neurologically confirmed MS according to the McDonald criteria ^[15]^ and excluding other neurological diseases (e.g., neuromyelitis optica and acute disseminated encephalomyelitis) were included in the study. The control group consisted of 25 age-matched females with no existing neurological disorders (EDSS = 0). Persons with known carotid artery disease, a previous medical history of diabetes mellitus and cardiovascular disease (CVD) were excluded in both groups. Disability assessments (EDSS) were available for 38 pwMS and biochemical and lifestyle data for 42 pwMS and the 25 controls.

### Ultrasound examination

The GE (General Electric) Logiq S8 and Logiq E9 ultrasound systems with Grey-scale imaging and Doppler facilities (colour and spectral Doppler) and a 9-12 MHz multifrequency linear transducer with coupling gel were used to assess the carotid arteries. A high frequency linear transducer was used to optimally visualise the superficial vasculature, namely the common carotid artery (CCA), internal carotid artery (ICA), external carotid artery (ECA) and vertebral artery (VA).

Grey-scale imaging was used to interrogate the major neck vessels for tortuosity, anatomical variation, plaque formation and measure the IMT of the CCA. The arterial wall is comprised of three layers (intima, media and adventitia) where intima-media thickening greater than 0.8 mm of the carotid arteries serves as a surrogate marker for early atherosclerosis. An automated average IMT, on the far side of the vessel wall, over a 2 cm segment of the mid CCA (3 cm proximal to the carotid bulb) was determined in the longitudinal plane (Figure 1). An IMT of 0.8 mm is regarded as the upper limit of normal ^[16]^.

**FIGURE 1.**
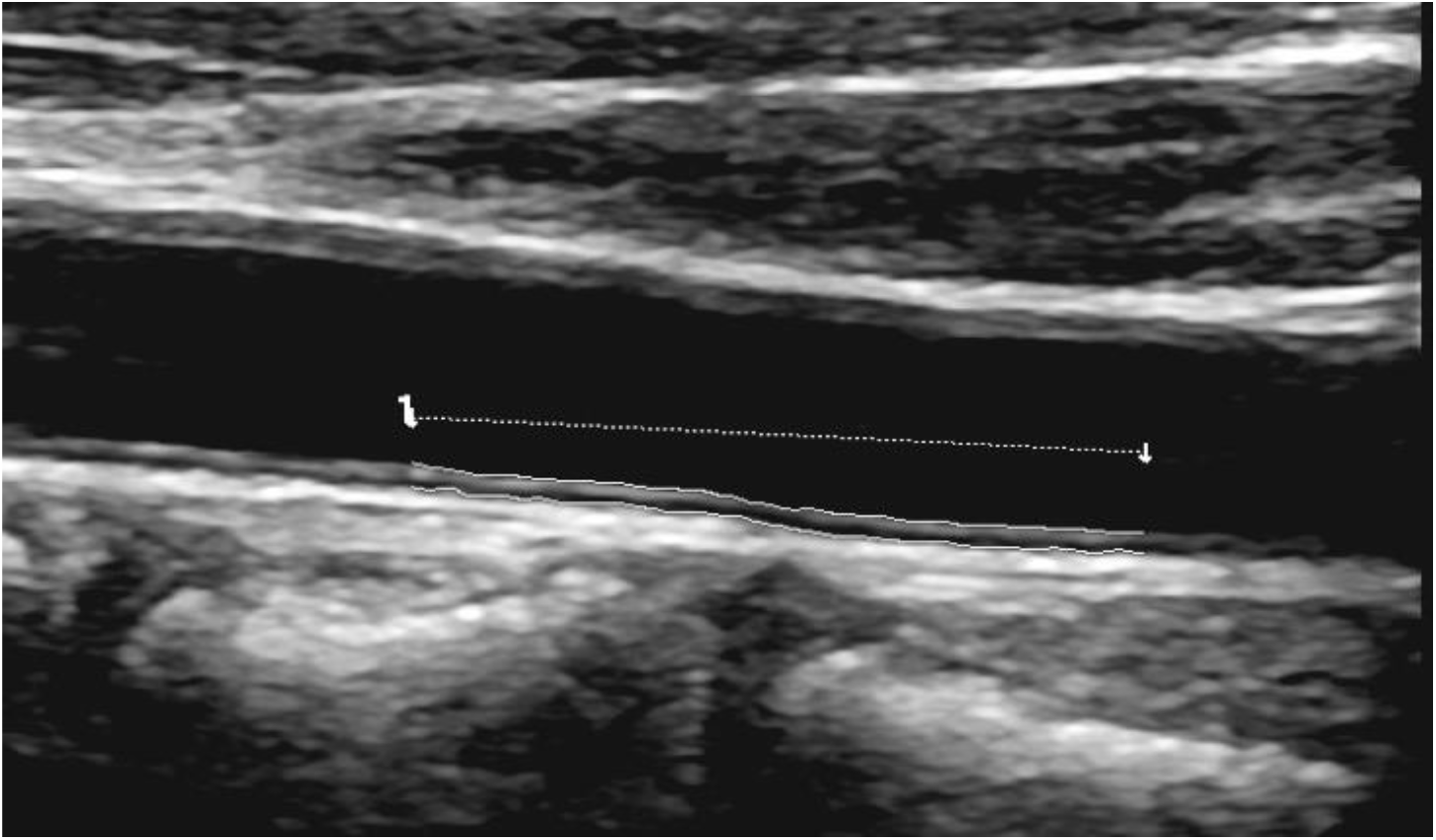
Grey-scale image of a normal left CCA in the longitudinal plane. Average IMT = 0.57mm (upper limit of normal = 0.8mm)

Colour Doppler was used to assess the vessels for patency, direction of blood flow, any colour-filling defects and presence of an occlusion. Spectral Doppler analysis was used to detect carotid artery stenosis by measuring the peak systolic velocity (PSV) and end-diastolic velocity (EDV) within the common carotid artery (CCA), internal carotid artery (ICA) external carotid artery (ECA) and vertebral artery (VA) as illustrated in Figure 2. A Doppler angle of 60° and a sample volume of 2 mm was used to produce an accurate velocity measurement with the angle cursor parallel to the wall of the segment of the vessel being sampled. The North American Symptomatic Carotid Endarterectomy Trials (NASCET) criteria were used to grade internal carotid artery stenosis, where a stenosis ≥ 70% is regarded as significant ^[17]^.

**FIGURE 2.**
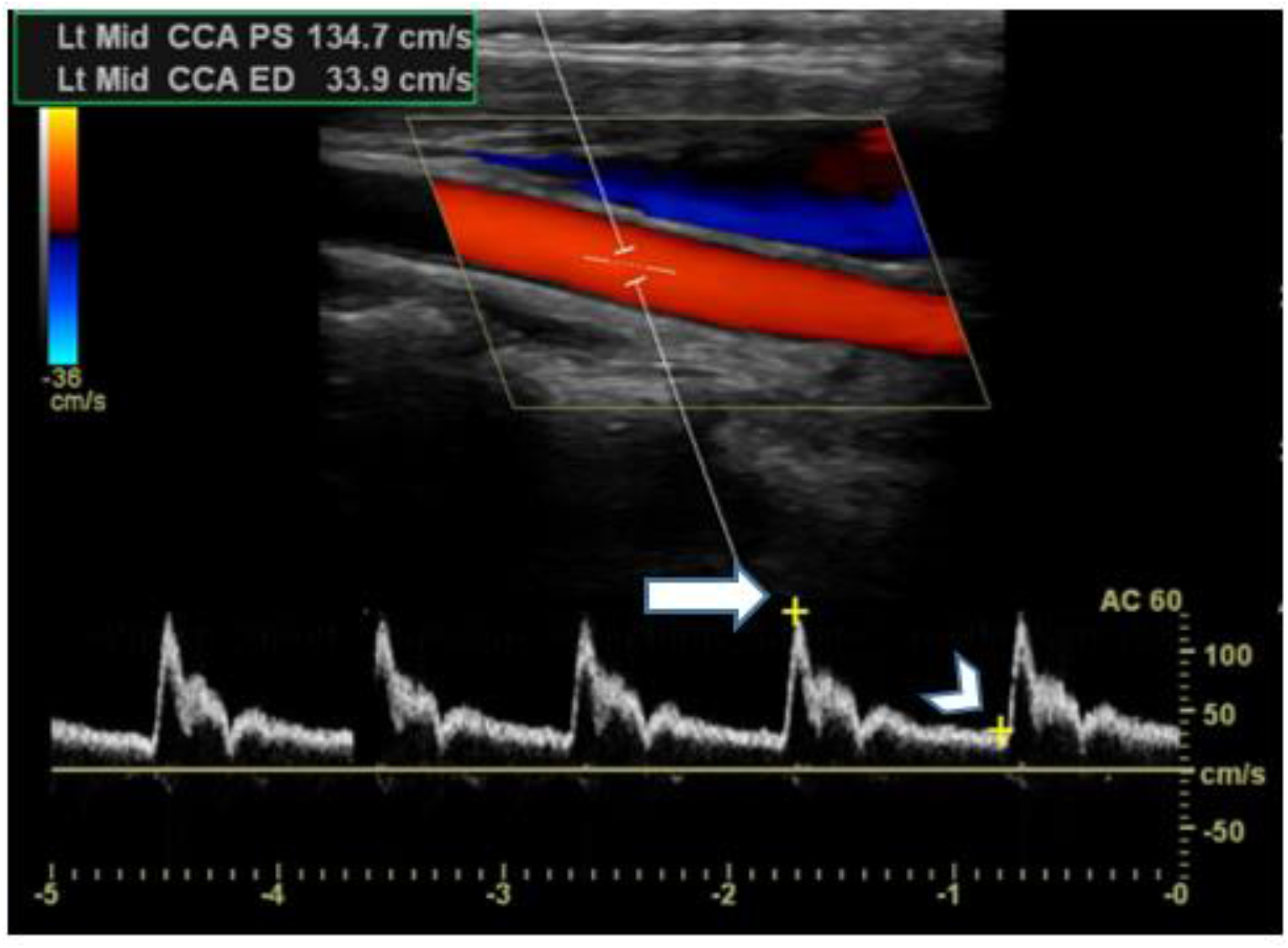
Colour Doppler and spectral Doppler analysis of a normal left common carotid artery (CCA) at a Doppler angle of 60°. Peak systolic velocity = 134.7 cm/s (solid arrow), end diastolic velocity = 33.9 cm/s (arrowhead). (Permission granted by patient #MS03)

#### Physical activity

Participants completed a medical and lifestyle questionnaire on entry to the study. Physical activity was self-reported and categorised into high (exercise four or more times a week), moderate (exercise two to three times a week) or low (exercise occasionally or complete lack of exercise).

### Biochemical analysis

Blood was drawn for biochemistry between 09h00 and 10h30 to standardise for diurnal variation as reported previously [18]. Laboratory analysis of the following cardiovascular-related biomarkers was performed by an accredited pathology laboratory: cholesterol, homocysteine, C-reactive protein (CRP), 25-OH vitamin D, serum folate and vitamin B12.

### Blood pressure measurements

Blood pressures were measured in right and left arms in 25 pwMS and 25 Controls.

### Disability measurement

The disability status was assessed by a clinician using the EDSS ^[13]^. The EDSS, which is regarded as the gold standard for measuring MS disability, ranges from 0 to 10, with higher scores indicating higher disability. For study purposes, the EDSS was assessed when the pwMS were in remission, so that their scores reflect the residual disability when not in a relapse.

### Comparison of Disability (EDSS) with Ultrasound parameters in pwMS

In order to assess whether ultrasound parameters were associated with disability, statistical comparisons were made between EDSS values, cIMT and blood flow parameters in the pwMS. Since cIMT is significantly associated with age ^[12]^ the relative effect of age was determined in the association between the cIMT and the EDSS using a regression summary for EDSS adjusted for IMT and Age.

### Statistical analysis

Statistical analyses were performed using Statistica™ version 13.4.0.14. Pearson’s correlations were used for testing relationships between disability (EDSS) and ultrasound measurements and adjustments for age and IMT were assessed using regression analysis (Table 2). Regression analysis was conducted to simultaneously investigate the IMT and age relationships with EDSS. For this regression, the normal probability plot was inspected to evaluate the normality assumption and was found to be acceptable. Regarding normality for Pearson correlations, corresponding non-parametric Spearman correlations were also calculated (not reported) and found to be similar to the Pearson correlations. Statistical significance was set as p < 0.05.

All tests were two-sided and various measures such as mean and standard deviation were used to describe the analyzed data. Mean is described as the average value of a group of numbers and standard deviation refers to how much variation there is within a group of values ^[19]^.

## Results

Table 1 provides the demographic and clinical characteristics of 51 pwMS and 25 controls.

**TABLE 1:**
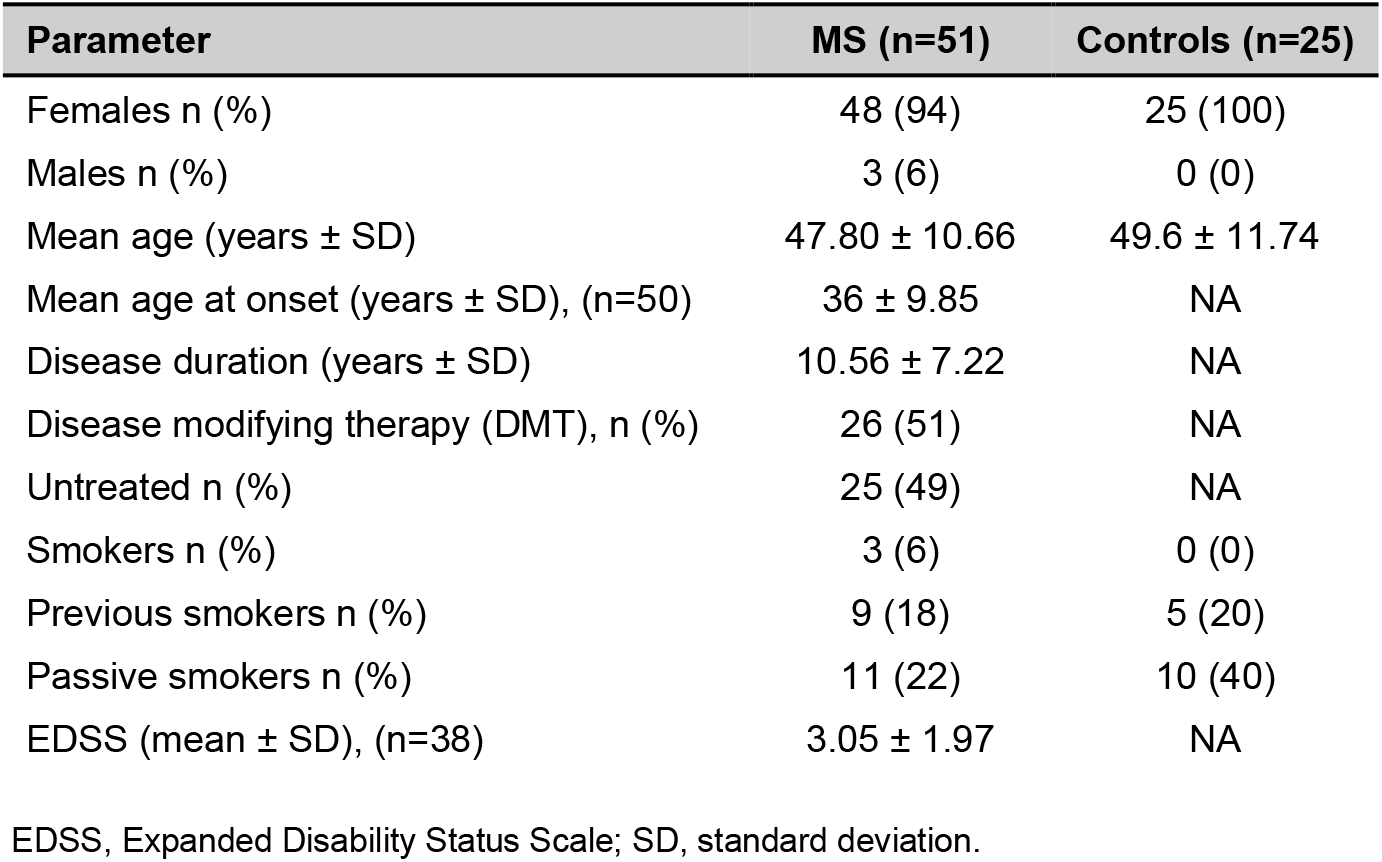
Characteristics of MS and control participants

| Parameter | MS (n=51) | Controls (n=25) |
| --- | --- | --- |
| Females n (%) | 48 (94) | 25 (100) |
| Males n (%) | 3 (6) | 0 (0) |
| Mean age (years ± SD) | 47.80 ± 10.66 | 49.6 ± 11.74 |
| Mean age at onset (years ± SD), (n=50) | 36 ± 9.85 | NA |
| Disease duration (years ± SD) | 10.56 ± 7.22 | NA |
| Disease modifying therapy (DMT), n (%) | 26 (51) | NA |
| Untreated n (%) | 25 (49) | NA |
| Smokers n (%) | 3 (6) | 0 (0) |
| Previous smokers n (%) | 9 (18) | 5 (20) |
| Passive smokers n (%) | 11 (22) | 10 (40) |
| EDSS (mean ± SD), (n=38) | 3.05 ± 1.97 | NA |

EDSS, Expanded Disability Status Scale; SD, standard deviation.

### Carotid and vertebral artery blood flow and carotid IMT

The right and left CCA, ICA, ECA and VA appeared patent and displayed normal blood flow velocities with no significant ICA stenosis (≥ 70 %) on grey-scale imaging, colour and spectral Doppler analyses. No significant difference in the right and left CCA, ICA and VA blood flow velocities and IMT was demonstrated between pwMS and controls. Three pwMS displayed an increased IMT of > 0.8 mm (0.9 - 1.0 mm).

### Association between IMT and EDSS

A statistically significant positive association was found between the EDSS and the right CCA IMT (r = 0.63; p < 0.001, Figure 3) and left CCA IMT (r = 0.49; p < 0.001). In the right CCA this association was already apparent at values between 0.4 - 0.7 mm, which are well below the 0.8 mm cut-off point for atherosclerosis, suggesting that even slight thickening of the IMT had an influence on disability. A significant association between the EDSS and age was also found (r = 0.49; p < 0.01) (Figure 3).

**Figure 3.**
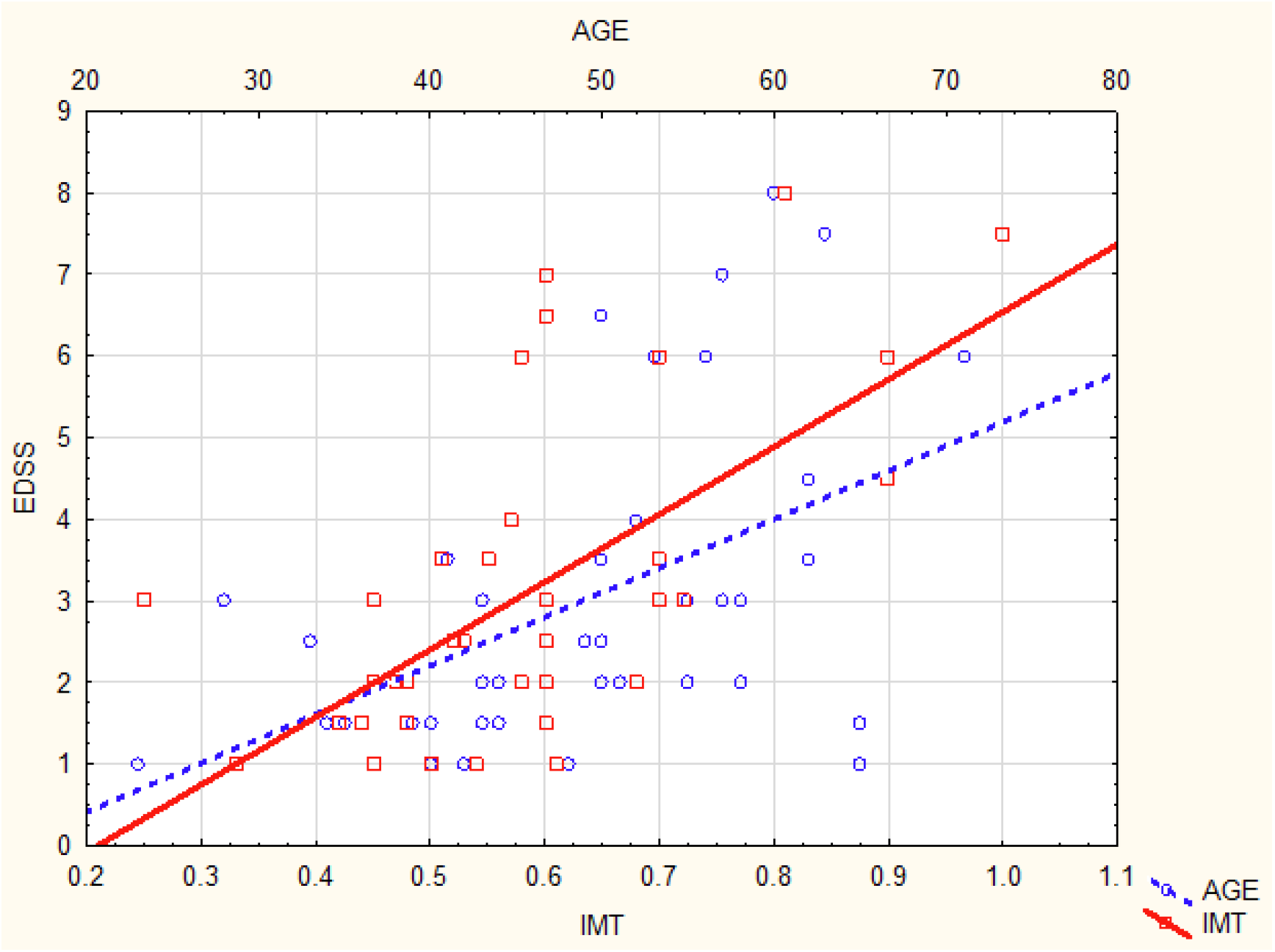
Significant associations between the EDSS, age and IMT of the RT CCA (*p*<0.001). Bottom axis denotes IMT and top axis denotes age. RT CCA, right common carotid artery; IMT, intima media thickness; EDSS, Expanded Disability Status Scale. An IMT above 0.8 mm is regarded as predictive of atherosclerosis risk.

#### Differential effects on EDSS after adjustment for IMT and Age

Both IMT and age showed significant associations with the EDSS. When IMT was adjusted for age in a regression analysis, the association between the EDSS and the IMT remained significant (p < 0.01). It is interesting to note that the age association was reduced to being significant only at 10% (p = 0.06, see Table 2).

**Table 2:** Regression Summary for EDSS adjusted for IMT and Age

| $n=38$, $R^2=0.46$ | Standardized coefficient | Standard error | $t$ (35) | $p$-value |
| --- | --- | --- | --- | --- |
| Age | 0.27(-0.004 – 0.64) | 0.14 | 1.95 | 0.06 |
| RT CCA IMT | 0.51 (0.24 – 0.78) | 0.14 | 3.70 | <0.01 |

RT=right; CCA=common carotid artery; IMT=intima media thickness

### Association between carotid and vertebral artery blood flow velocities and disability

Carotid and vertebral artery PSV (arterial blood flow velocity in systole) and EDV (arterial blood flow velocity in diastole) were inversely associated with disability in pwMS. The EDSS was significantly negatively associated with the PSV of the left CCA (r = -0.40; p = 0.01), left ICA (r= -0.42; p = 0.01), right VA (r = -0.42; p < 0.001, Figure 4) and the EDV of the left ICA (r = -0.47; p < 0.001, Figure 5).

**FIGURE 4.**
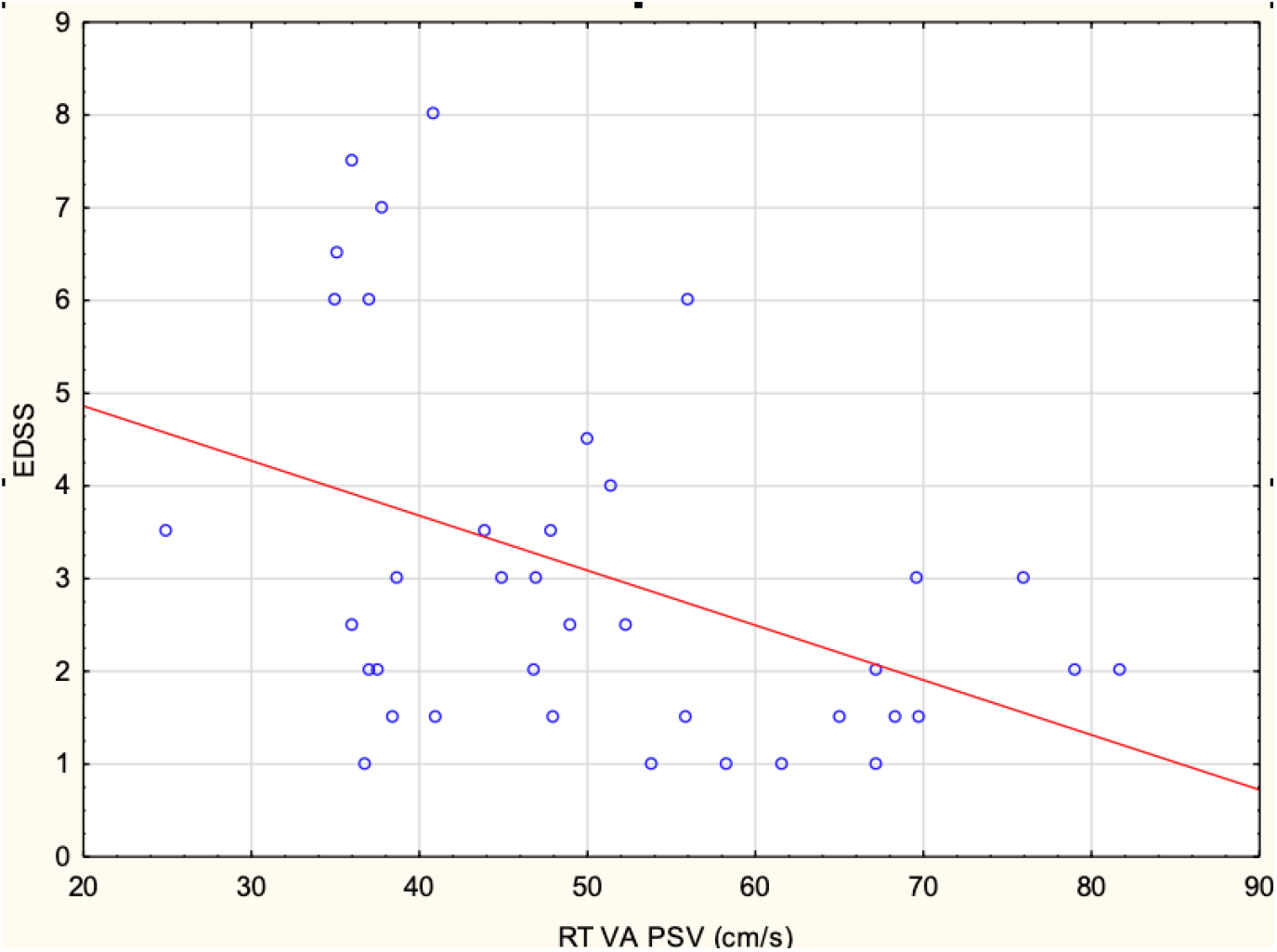
Significant negative associations between EDSS and the PSV of the RT VA. EDSS, Expanded Disability Status Scale; RT VA, right vertebral artery; PSV, peak systolic velocity. Normal PSV is ∼40-60cm/s.

**FIGURE 5.**
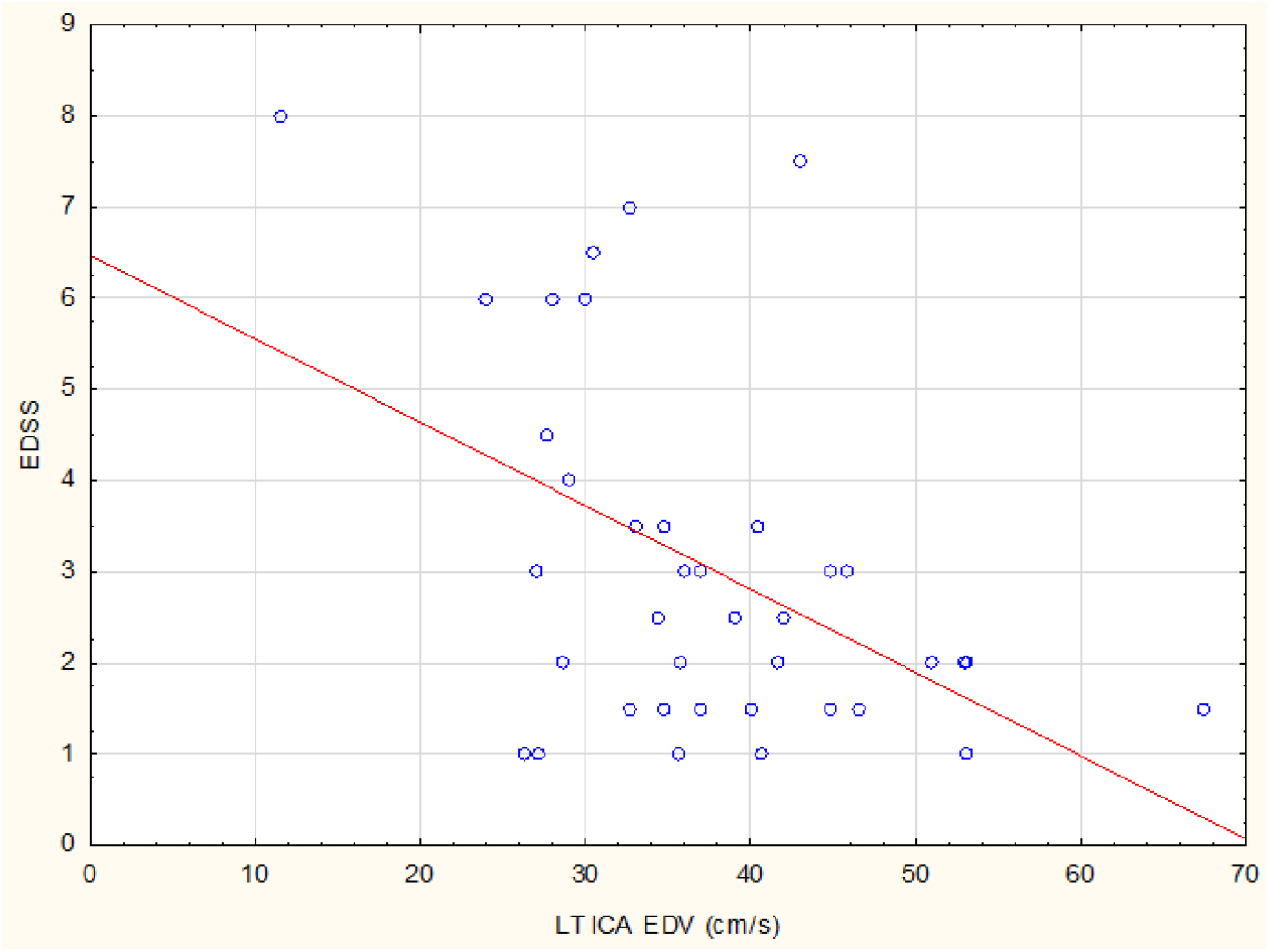
Significant negative associations between EDSS and the EDV of the LT ICA EDSS, Expanded Disability Status Scale; LT ICA, left internal carotid artery; EDV, end-diastolic velocity. Normal EDV is ∼40cm/s.

#### Association between carotid and vertebral artery blood flow velocities and lifestyle parameters in the MS cohort

Significant associations were found between ultrasound parameters related to blood flow and biochemistry/lifestyle factors in the pwMS.

Improved associations (positive r-values) between PSV and biochemical factors were found for higher serum folate and vitamin D concentrations and greater physical activity, while increased blood levels of homocysteine and cholesterol had worse associations (negative r-values) with PSV (p ≤ 0.01; Table 3).

**Table 3.** Significant positive and negative associations between blood flow parameters and biochemistry/lifestyle factors in MS (n=42)

| Ultrasound parameter | Biochemical/Lifestyle factors | Pearson |  |
| --- | --- | --- | --- |
|  |  | r | p |
| PEAK SYSTOLIC VELOCITY (PSV) |  |  |  |
| RT VA PSV (cm/s) | Serum folate | 0.43 (0.14, 0.65) | <0.01 |
| LT VA PSV (cm/s) | Vitamin D | 0.41 (0.11, 0.64) | <0.01 |
| RT VA PSV (cm/s) | Physical activity | 0.34 (0.03, 0.59) | 0.03 |
| LT ICA PSV (cm/s) | Physical activity | 0.40 (0.10, 0.63) | 0.01 |
| RT CCA PSV (cm/s) | Homocysteine | -0.46 (-0.67, -0.18) | <0.01 |
| RT ECA PSV (cm/s) | Homocysteine | -0.33 (-0.58, -0.02) | 0.04 |
| LT VA PSV (cm/s) | Cholesterol | -0.46 (-0.67, -0.18) | <0.01 |
| END DIASTOLIC VELOCITY (EDV) |  |  |  |
| RT CCA EDV (cm/s) | Cholesterol | -0.39 (-0.62, -0.10) | 0.01 |
| LT CCA EDV (cm/s) | Cholesterol | -0.37 (-0.61, -0.07) | 0.02 |

LT = left; RT= right; PSV = peak systolic velocity; EDV = end-diastolic velocity; ICA = internal carotid artery; CCA = common carotid artery; VA = vertebral artery; PROX = proximal; ECA = external carotid artery;

Other associations (p < 0.05) were the following: Physical activity was associated with lower left CCA IMT (r = -0.37; p = 0.02), higher PSV of the proximal left ECA (r = 0.34; p = 0.03) and higher EDV of the left ICA (r = 0.37; p = 0.02).

Serum vitamin D was associated with higher PSV of the right CCA (r = 0.37; p = 0.02) and left CCA (r = 0.35; p = 0.03). CRP, an indicator of inflammation, was associated with lower EDV of the left ICA (r = -0.32; p = 0.04).

#### Association between ultrasound parameters, smoking and disease modifying therapy (DMT) in the MS cohort

Smokers were assessed as active smokers, previous smokers and passive smokers. When the three groups were combined in the MS cohort, smoking significantly (p = 0.01) decreased the EDV of the left VA compared to no smoking. In passive smokers, there were non-significant trends for lower EDV of the left VA, the ICA and the right VA compared to non-smokers (p=0.07, 0.09 and 0.08 respectively). In the cohort of 51 pwMS, 26 were treated with disease modifying therapy (DMT) and 25 were not (Table 1). There was no association between DMT use and the EDSS (p = 0.56); however, people on DMT had a higher EDV of the left CCA (p < 0.01) than those not on DMT.

#### Association between ultrasound parameters and blood pressure in the MS and Control cohorts

There were no associations between blood pressure (BP) and EDSS in the pwMS. BP results were available for the right arms and the left arms of 25 MS and 25 control participants. Right arm BPs were consistently higher than left arm BPs in both cohorts but not significantly. MS BPs were higher than those of controls, but not significantly (Table 4).

**Table 4.**
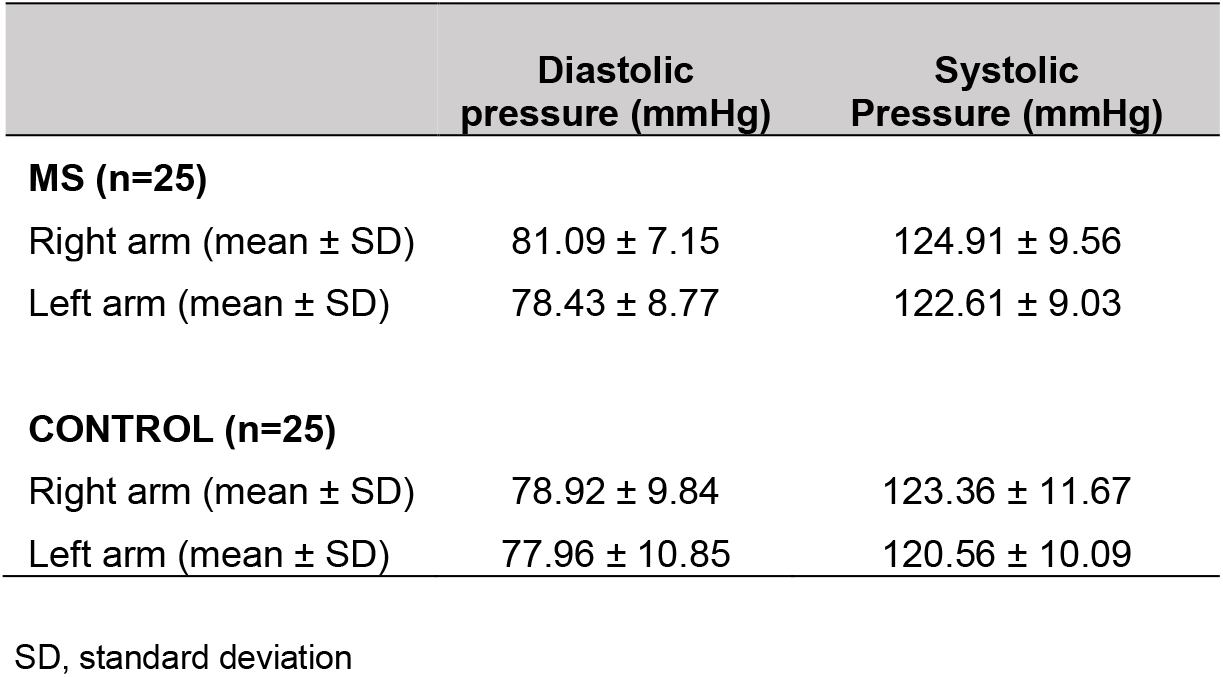
Blood pressures in MS and control participants

Unexpectedly, there were no significant associations between BP, cIMT and arterial blood flow parameters in the pwMS, while in contrast, several significant associations were seen in the control group (Table 5).

**Table 5.**
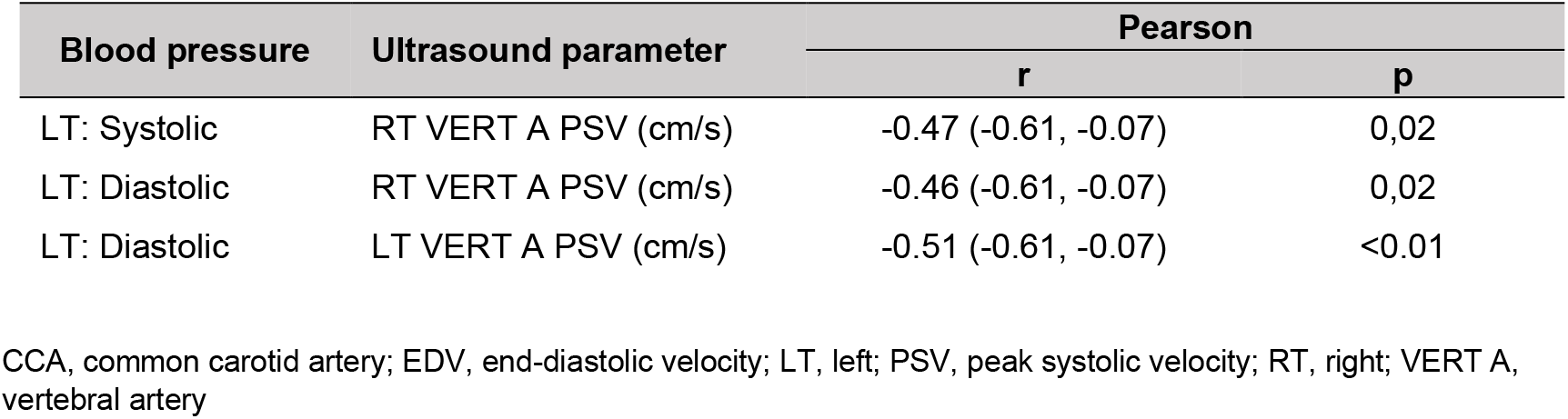
Associations between blood pressure and blood flow parameters in control participants

| Blood pressure | Ultrasound parameter | Pearson |  |
| --- | --- | --- | --- |
|  |  | r | p |
| LT: Systolic | RT VERT A PSV (cm/s) | -0.47 (-0.61, -0.07) | 0,02 |
| LT: Diastolic | RT VERT A PSV (cm/s) | -0.46 (-0.61, -0.07) | 0,02 |
| LT: Diastolic | LT VERT A PSV (cm/s) | -0.51 (-0.61, -0.07) | <0.01 |

CCA, common carotid artery; EDV, end-diastolic velocity; LT, left; PSV, peak systolic velocity; RT, right; VERT A, vertebral artery

## Discussion

The present study found that MS disability (EDSS) was significantly associated with vascular parameters, including increased cIMT as well as reduced blood flow velocities within the carotid and vertebral arteries, suggesting that vascular perfusion had a profound influence on disability, confirming the results of previous studies ^[10,20,21]^. Age was also significantly associated with the EDSS, but after adjustment for cIMT, age lost its significance (p=0.06; Table 2). In contrast, when cIMT was adjusted for age, the association between cIMT and EDSS remained significant (p < 0.01). This may suggest that factors that can be controlled by the individual, such as lifestyle decisions, may override factors that cannot be modified, such as age, in the outcome of disability progression. We found that cIMT values were highly significantly associated with EDSS scores (r = 0.63, p = 0.001) and that this association was already apparent at values between 0.4 - 0.7 mm, which are well below the 0.8 mm cut-off point for atherosclerosis (Figure 3), confirming the result of a previous study by Nelson et al. ^[22]^. The associations between MS disability and ultrasound imaging is currently under-investigated, therefore literature on associations between the EDSS and IMT and extracranial arterial blood flow remains limited ^[22,23]^. However, an increasing number of studies have confirmed an important role of the vasculature in MS disability progression ^[10,20,21]^.

The explanation for these findings may be that a healthy vasculature provides the brain with the oxygen and nutrients required for optimal myelin synthesis, while atherosclerotic vasculature may impede blood flow. Epidemiological studies suggest that pwMS are at a higher risk for stroke and atherosclerosis ^[7]^. Atherosclerosis is described as an inflammatory disease of the vascular wall (usually arterial), with the deposition of oxidised lipoproteins (LDL-cholesterol) and the participation of the immune system ^[24]^. The process is initiated by endothelial dysfunction, due to damage caused by elevated LDL-cholesterol and homocysteine, diabetes, hypertension, and free radicals from, for example, cigarette smoke, viruses or bacteria. The endothelium lines all blood vessels and in normal physiological conditions, contributes to vascular homeostasis by actively regulating vascular tone, controlling permeability between the bloodstream and the underlying vascular wall, regulating medial smooth muscle cell growth and controlling platelet function, coagulation and fibrinolysis ^[25]^. Endothelial damage increases the adhesiveness and permeability of the endothelium, allowing entry of monocytes and T-lymphocytes into the vessel wall ^[26]^. LDL particles undergo progressive oxidation and are internalised by macrophages, resulting in the formation of vascular plaques that may, in advanced cases, completely occlude the blood vessels ^[27]^. It was also demonstrated that the Framingham General Cardiovascular Disease Risk Score (FR) was significantly associated with the EDSS (p < 0.001) ^[28]^. Valavanis et al. ^[23]^ found that an ultrasound index of atherosclerosis (ATHUS score) based on arterial stenosis and the IMT of the carotid and femoral arteries, was significantly associated with cognitive function and the EDSS.

Associations have also previously been demonstrated between CVD and systolic interarm BP differences of more than 5 mmHg ^[9]^. In the present study, BPs were higher in pwMS than controls, but not significantly, while non-significant interarm differences in systolic BP were found in both the MS and control cohorts (Table 4). Unexpectedly, BP measurements in the pwMS were not associated with cIMT or blood flow parameters, while in contrast, significant associations were found for these parameters in the controls. Elucidating the reasons for these differences is beyond the scope of this study and future work is warranted to substantiate these findings.

The present study demonstrated that the presence of biochemical markers in the blood significantly enhanced or impeded blood flow in the pwMS (Table 2). As would be expected, cholesterol, homocysteine and CRP, a marker of inflammation with direct proinflammatory effects, were negatively associated with blood flow, while the nutrients folate and vitamin D were positively associated with blood flow parameters. Folate, found in fruits and vegetables, is essential for lowering homocysteine. These results confirm previous findings of Davis et al. ^[18]^ who showed that cholesterol correlated with homocysteine levels, which were in turn significantly lowered by higher folate in the diet, and that intake of fruits and vegetables was significantly associated with improved EDSS in pwMS. Fruits and vegetables also contain antioxidants, which reduce free-radical formation by modified LDL ^[29]^.

Different dietary patterns have shown associations with cIMT ^[30]^. An ongoing prospective, randomized, single-blind, controlled trial demonstrated that a Mediterranean diet rich in extra virgin olive oil significantly (p < 0.001) decreased the cIMT over five years, maintained at seven years, in 1002 coronary heart disease patients, with ultrasound assessments ^[30]^. Conversely, diets high in saturated fat, trans fatty acids and dietary cholesterol, and low in unsaturated fats, are positively associated with cIMT progression. Another randomised clinical trial found that adherence to the Mediterranean diet was significantly inversely associated with the EDSS in pwMS ^[4]^. Other studies have confirmed the role of diet in protecting the vascular system and brain against oxidation and inflammation ^[26]^.

Furthermore, the present study found that blood flow parameters were significantly associated with disability in pwMS, confirming previous studies ^[22,20,21]^. Carotid and vertebral artery PSV (arterial blood flow velocity in systole) and EDV (arterial blood flow velocity in diastole) were inversely associated with disability in pwMS. The ICA EDV showed an inverse relationship with the EDSS (Figure 5), while the lowest EDSS values were found between optimal PSV 37 and 68 cm/s (Figure 4). These associations confirm that increased MS disability may be related to abnormal blood flow to the brain due to cerebral hypoperfusion ^[2]^ demonstrating a reduction in cerebral blood flow in pwMS which is associated with hypoxia leading to white matter damage. Oligodendrocytes are extremely vulnerable to a hypoxic environment, resulting in demyelination and an increase in formation of white matter lesions ^[1,2]^.

The present study also identified opposite effects of smoking (active and passive), and physical activity on blood flow parameters. These results confirm previous findings of Davis et al. ^[18]^ that the EDSS was higher in smokers than in non-smokers in pwMS.

## Conclusion

This study confirmed that vascular factors play an integral part in the pathophysiological processes associated with MS, demonstrating by vascular ultrasound the significant association between cIMT and MS disability at values of cIMT even lower than the cut-off for atherosclerosis. Since the cIMT is a reflection of vessel wall patency in the brain, this finding provides important information for disease management of people diagnosed with MS, suggesting that modification of their diet and lifestyle may promote the unhindered flow of essential factors (oxygen and nutrients) through the BBB into the brain that are needed for the maintenance of oligodendrocytes and myelin production. If this finding is confirmed by further studies it may lead to a better understanding of disability prevention in MS.

## Data Availability

All data produced in the present study are available upon reasonable request to the authors

## Acknowledgements

We acknowledge Drs Bergman, Ross and partners Radiologists for allowing us to perform the research within their Ultrasound department. We also thank all study participants and the MS Care Trust, Cape Town, South Africa.

## Informed Consent

This investigation was a collaborative study between two universities in the Western Cape, South Africa. Ethical approval was granted by the Faculty of Health and Wellness Sciences Research Ethics Committee (FREC) (Reference no: CPUT/HW-REC 2017/H4) and the Human Research Ethics Committee (HREC) of the University of Stellenbosch (Reference no: N07/09/203). All study participants signed informed consent for their data to be used in the study. Ultrasound examinations of these participants were performed, by permission, at a private radiology practice.

